# Spatial Recurrence Is More Common Than Local Recurrence in Youth Athletes With Groin, Anterior Knee, and Heel Pain: A Retrospective Observational Study

**DOI:** 10.64898/2026.02.04.26345530

**Authors:** Shinsuke Sakoda

**Affiliations:** Department of Sports Medicine, Ashiya Central Hospital, Fukuoka, Japan

**Keywords:** Youth athletes, musculoskeletal pain, recurrence, spatial recurrence, groin pain, anterior knee pain, heel pain

## Abstract

**Objective:** To determine the incidence and transition patterns of local and spatial recurrence among youth athletes presenting with groin pain (GP), anterior knee pain (AKP), and heel pain (HP).

**Design:** Retrospective observational study.

**Setting:** Single sports medicine clinic.

**Participants:** Youth athletes who visited the clinic between January 2017 and November 2025 for GP, AKP, or HP. A total of 769 clinical episodes were included.

**Independent Variables:** Pain-site transitions and laterality were extracted from electronic medical records. No therapeutic interventions were evaluated. Recurrence patterns were classified as local or spatial and further subclassified into contralateral same-site, adjacent-site, and remote-site recurrence.

**Main Outcome Measure:** True recurrence, defined as an episode with at least one prior visit for GP, AKP, or HP within the preceding 12 months.

**Results:** Among 769 episodes, 130 episodes (16.9%; 95% confidence interval [CI], 14.4– 19.7) represented true recurrence. Local recurrence accounted for 57 episodes (7.4%; 95% CI, 5.8–9.5), whereas spatial recurrence accounted for 73 episodes (9.5%; 95% CI, 7.6–11.8). Among spatial recurrences, 26 episodes (35.6%) were contralateral same-site, 42 (57.5%) adjacent-site, and 5 (6.8%) remote-site recurrence. Transitions most frequently occurred between GP and AKP and between AKP and HP, whereas direct transitions between GP and HP were uncommon.

**Conclusions:** Recurrence after symptom resolution in youth athletes more often involves spatial transitions to anatomically adjacent sites rather than simple local recurrence. These findings support interpreting recurrence within a whole-body functional framework and may facilitate refined recurrence risk assessment and comprehensive intervention strategies.

**Clinical Relevance:** Clinicians should evaluate recurrent pain beyond the symptomatic region, which may improve recurrence prevention and return-to-sport decision-making.

## Introduction

Sports-related pain in adolescent athletes is one of the most common chief complaints encountered in sports medicine clinics. In particular, groin pain (GP), anterior knee pain (AKP), and heel pain (HP) are representative non-traumatic pain syndromes frequently observed in daily clinical practice and are thought to be associated with growth-specific anatomical characteristics, including open physes and the vulnerability of musculotendinous attachment sites^1–3^.

Numerous epidemiological studies have reported the incidence, prevalence, and persistence of these pain conditions. For example, calcaneal apophysitis has been reported to recur or persist in a substantial proportion of patients, and AKP may persist into late adolescence in a considerable number of cases^1, 4^. However, in many previous studies, persistent symptoms and true recurrence after a symptom-free interval have not been clearly distinguished and have often been treated as the same outcome^3–5^.

Moreover, most existing studies have focused on a single anatomical site or a single diagnosis, and few have systematically evaluated whether pain in adolescent athletes reappears at the same anatomical site or shifts to a different site over time, that is, the inter-site patterns of pain recurrence. In clinical practice, it is not uncommon to encounter patients who develop pain at a different site after initial symptom resolution and subsequently return for medical care; however, this phenomenon has not been sufficiently quantified or conceptually structured^6, 12^.

Adolescence is characterized by concurrent changes in skeletal maturation, muscle– tendon flexibility, and training volume and intensity, which may result in transient imbalances in tissue strength between the musculotendinous unit and its bony attachment. Such biomechanical dynamics may not be adequately explained by conventional models that conceptualize pain solely as a localized pathology at a single anatomical site^10–12^.

Therefore, the purpose of this study was to evaluate revisit patterns in adolescent athletes with GP, AKP, and HP by defining recurrence based on visit history and classifying revisit patterns into local recurrence and spatial recurrence. By recharacterizing sports-related pain in adolescents not merely as recurrence at a single site but as a dynamic pattern across multiple anatomical sites, this study aims to provide fundamental insights that may contribute to improved pain assessment and the development of more effective recurrence-prevention strategies in adolescent athletes.

## Methods

### Study Design and Participants

This was a single-center retrospective observational study of adolescent athletes who visited the sports medicine clinic at our institution between January 2017 and November 2025.

### Unit of Analysis and Episode Definition

Each clinic visit was counted as one episode. Revisit episodes were identified by linking patient records using unique patient identification numbers in the electronic medical record system. Only visits to the sports medicine clinic at our institution were included in the analysis.

### Exclusion Criteria

Episodes involving acute traumatic injuries, including sprains, dislocations, and fractures; injuries unrelated to sports activities, such as those occurring during daily activities or traffic accidents; postoperative conditions; and bacterial infections were excluded from the analysis.

### Study Population

All included patients had open physes at the time of their initial visit. After applying the exclusion criteria, a total of 1,359 episodes (mean age, 11.8 ± 2.0 years; 254 females) were included in the final analysis.

### Pain Categories

Chief complaints were categorized into three groups: GP, AKP, and HP.

### Outcome Definitions and Classification of Revisit Patterns

The primary outcome was the proportion of episodes classified as true recurrence among episodes with GP, AKP, or HP. True recurrence was operationally defined as a revisit episode in which the interval between index visits for any of these pain categories was less than 12 months; episodes separated by 12 months or longer were classified as independent (new) episodes.

The 12-month reference period was selected because physical maturation and training environments in adolescent athletes typically change on an annual basis, making recurrence assessment under comparable biological and exposure conditions less reliable over longer intervals.

True recurrence was classified into two categories based on the pain site and laterality at revisit:

1. Local recurrence: recurrence at the same pain site on the same side.
2. Spatial recurrence: recurrence occurring either at the same anatomical site on the contralateral side or at a different pain site.

Spatial recurrence was further subclassified into three groups based on the anatomical relationship between pain sites:

1. Contralateral same-site recurrence: recurrence at the same pain site on the opposite side.
2. Adjacent-site recurrence: transitions between anatomically adjacent pain sites (GP ↔ AKP and AKP ↔ HP).
3. Remote-site recurrence: transitions between non-adjacent pain sites (GP ↔ HP). For each subclassification, the number of episodes and the directional frequency of transitions between pain sites were summarized.

### Statistical Analysis

Statistical analyses were performed using descriptive statistics. The proportions of local and spatial recurrence were expressed as percentages of true recurrence episodes, with 95% confidence intervals calculated using the Wilson score method. Subclassifications of spatial recurrence and inter-site transitions were summarized descriptively.

### Ethical Considerations

This study was approved by the institutional review board of our institution. Because of the retrospective study design, informed consent was obtained using an opt-out approach.

### Use of Artificial Intelligence

Generative artificial intelligence (ChatGPT, OpenAI, San Francisco, CA, USA) was used solely to assist with English language editing and clarity of expression. All study design, data collection, statistical analyses, interpretation of results, and final manuscript content were performed and verified by the authors.

## Results

### Distribution of Pain Categories

Among the 1,359 episodes included in the final analysis, 769 episodes were classified into the three pain categories: GP, AKP, and HP. The distribution was 157 GP episodes (11.6%), 396 AKP episodes (29.1%), and 216 HP episodes (15.9%).

### Age and Sex Distribution

The mean age differed significantly among the three pain categories (GP, 12.5 ± 2.2 years; AKP, 12.0 ± 1.7 years; HP, 10.9 ± 1.7 years; one-way analysis of variance, p < 0.001). The proportion of female patients was 15.9% (25/157) in the GP group, 18.9% (75/396) in the AKP group, and 17.1% (37/216) in the HP group, with no significant difference among the groups (chi-square test, not significant).

### Revisit Patterns

Among the 769 episodes with GP, AKP, or HP, true recurrence—defined as a revisit episode with a history of at least one visit for any of the three pain categories within the preceding 12 months—was observed in 130 episodes (16.9%; 95% confidence interval [CI], 14.4%–19.7%).

Of these true recurrence episodes, 57 episodes (7.4%; 95% CI, 5.8%–9.5%) were classified as local recurrence, and 73 episodes (9.5%; 95% CI, 7.6%–11.8%) were classified as spatial recurrence. The proportions of local recurrence by pain category were 3.8% (6/157) for GP, 10.4% (41/396) for AKP, and 4.6% (10/216) for HP.

Among the 73 spatial recurrence episodes, 26 episodes (35.6%; 95% CI, 25.6%– 47.1%) were classified as contralateral same-site recurrence, 42 episodes (57.5%; 95% CI, 46.1%–68.2%) as adjacent-site recurrence, and 5 episodes (6.8%; 95% CI, 3.0%–15.1%) as remote-site recurrence, as summarized in Table 1.

**Table 1.** Distribution of Local and Spatial Recurrence Patterns Among Anatomically Related Pain Sites in Youth Athletes. Footnote Percentages for overall recurrence, local recurrence, and spatial recurrence are calculated using the total number of episodes (n = 769) as the denominator. Percentages for subcategories of spatial recurrence are calculated using the number of spatial recurrence episodes (n = 73) as the denominator. CI indicates confidence interval.

| Category | Cases (n) | % (95%CI) |
| --- | --- | --- |
| Total episodes | 769 |  |
| True recurrence | 130 | 16.9 (14.4–19.7) |
| –Local recurrence | 57 | 7.4 (5.8–9.5) |
| –Spatial recurrence | 73 | 9.5 (7.6–11.8) |
| –Contralateral same-site | 26 | 35.6 (25.6–47.1) |
| –Adjacent-site | 42 | 57.5 (46.1–68.2) |
| –Remote-site | 5 | 6.8 (3.0–15.1) |

Contralateral same-site recurrence occurred in 11 GP episodes, 10 AKP episodes, and 5 HP episodes. Adjacent-site recurrence included 7 transitions from GP to AKP and 9 transitions from AKP to GP, as well as 11 transitions from AKP to HP and 15 transitions from HP to AKP. In contrast, remote-site recurrence was rare, with 4 transitions from GP to HP and 1 transition from HP to GP, for a total of 5 episodes (Figure 1).

**Figure 1.**
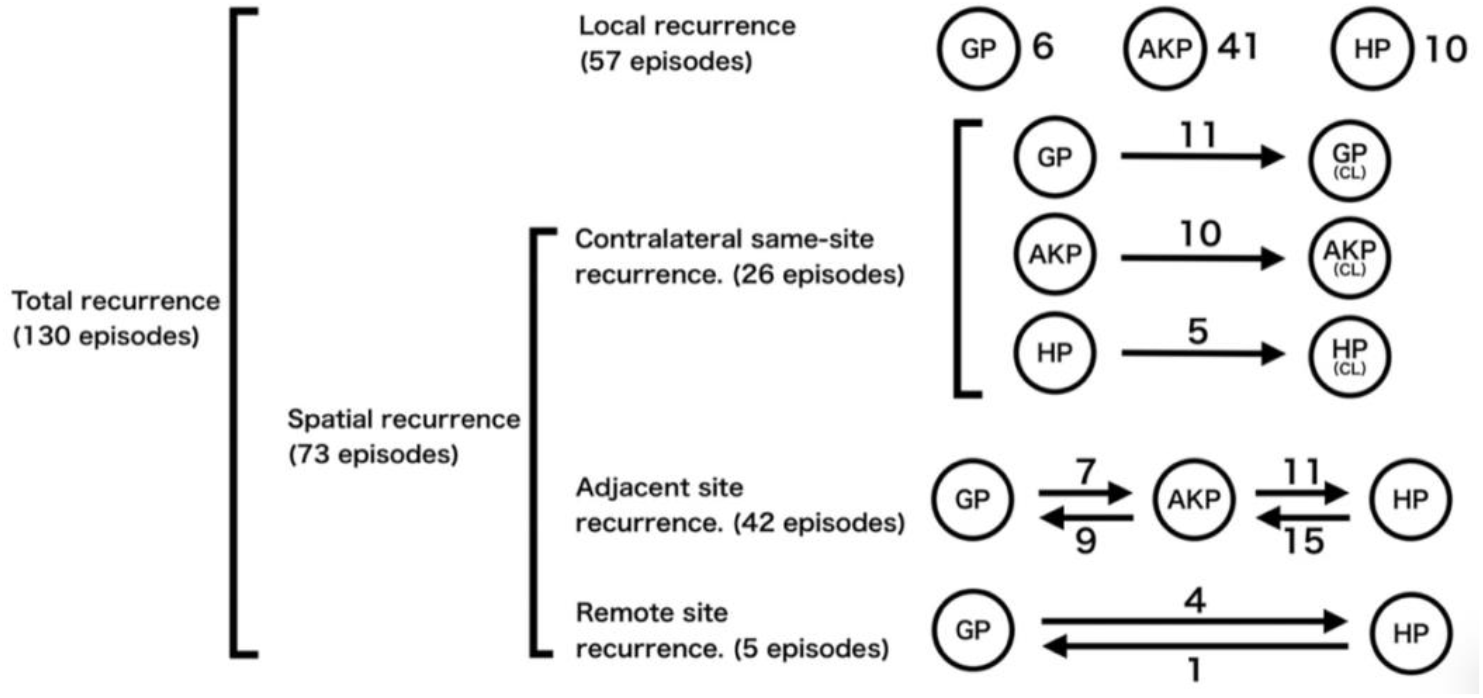
Patterns of local and spatial recurrence among groin pain (GP), anterior knee pain (AKP), and heel pain (HP). Among 130 recurrent episodes, 57 were classified as local recurrence (same side and same pain site), and 73 were classified as spatial recurrence. Spatial recurrence was further subdivided into contralateral same-site recurrence (n = 26), adjacent-site recurrence (n = 42), and remote-site recurrence (n = 5). Arrows indicate the direction of transitions between pain sites, and the numbers adjacent to each arrow represent the number of observed transition episodes. Contralateral same-site recurrence refers to recurrence at the same anatomical site on the opposite side (CL). Adjacent-site recurrence refers to transitions between anatomically adjacent regions (GP ↔ AKP and AKP ↔ HP), whereas remote-site recurrence refers to transitions between non-adjacent regions (GP ↔ HP). Abbreviations: GP, groin pain; AKP, anterior knee pain; HP, heel pain; CL, contralateral.

## Discussion

Numerous studies have reported the incidence and persistence of sports-related pain in adolescent athletes, including HP and AKP. Calcaneal apophysitis has been shown to recur or persist in a substantial proportion of patients, and AKP may persist into late adolescence in many individuals. However, in much of the existing literature, persistent symptoms and true recurrence after a symptom-free interval have not been clearly distinguished, and few studies have systematically examined whether recurrence occurs at the same anatomical site or shifts to a different site^3–6^.

A key feature of the present study is that revisit episodes were defined using an objective criterion based on whether a visit for any of the three pain categories (GP, AKP, or HP) occurred within the preceding 12 months, and revisit patterns were structurally classified into local and spatial recurrence. Using this approach, the revisit rate for the same pain category was relatively low (approximately 4–10%), whereas the overall proportion of true recurrence across the three pain categories reached 16.9%. These findings suggest that sports-related pain in adolescence may be more accurately characterized as a dynamic clinical pattern involving transitions across anatomical sites rather than as chronic persistence at a single site^1, 4, 5^. To our knowledge, this is the first study to quantitatively visualize inter-site recurrence patterns across multiple pain sites in adolescent athletes, representing a key element of the novelty of this work.

The three pain conditions examined in this study share common anatomical features, as they occur at sites with apophyses that are vulnerable to traction stress during growth. Adolescence is characterized by the coexistence of open physes, progressive physical maturation, changes in muscle–tendon flexibility, and fluctuations in training volume and intensity, which can create transient imbalances in tissue strength between the musculotendinous unit and its bony attachment^10, 11^. As a result, pain may not repeatedly manifest at the same anatomical site but may instead shift over time toward apophyseal regions with relatively lower load tolerance.

Consistent with this interpretation, most spatial recurrences in the present study occurred between anatomically adjacent sites, suggesting that mechanical load may be redistributed across neighboring segments within the kinetic chain. Such revisit patterns highlight the limitations of conventional models that conceptualize pain solely as a localized pathology at a single site and support a broader biomechanical and systems-based interpretation of injury mechanisms^7, 9, 12^.

Beyond a site-specific paradigm, these findings support a conceptual shift toward viewing adolescent sports-related pain as a network-level phenomenon rather than a localized pathology. Traditional models implicitly assume that recurrence reflects persistent vulnerability at a single anatomical site, leading to predominantly site-focused diagnostic and therapeutic strategies. In contrast, the present data demonstrate that symptom recurrence frequently involves transitions across anatomically related regions, suggesting dynamic redistribution of mechanical load within the growing musculoskeletal system^12^.

We propose the concept of an “apophyseal vulnerability network,” in which multiple growth-related vulnerable sites function as interconnected nodes within a kinetic chain. Changes in growth velocity, tissue maturation, training load, and movement patterns may alter relative load tolerance among these nodes over time, resulting in shifting manifestations of pain rather than repeated failure at a single site. This framework provides a systems-based perspective for interpreting recurrent pain during adolescence and offers a theoretical basis for integrating injury surveillance, biomechanical assessment, and longitudinal load management.

From a clinical perspective, symptom resolution at a single anatomical site should not necessarily be interpreted as restoration of the underlying functional capacity in adolescent athletes. Sports-related pain during growth may reflect a mismatch between rapidly increasing external load demands and the still-maturing global neuromusculoskeletal system. Even when local symptoms improve, insufficient recovery or adaptation of global function may predispose athletes to recurrent pain either at the same site or at other anatomically or functionally related regions.

Accordingly, clinical management should extend beyond localized treatment and incorporate sufficient patient education, comprehensive functional assessment, and interventions aimed at improving global movement capacity, load tolerance, and recovery strategies^7, 8^. In follow-up after symptom resolution, priority should be placed on comprehensive evaluation and proactive enhancement of global function. The goals extend beyond reducing recurrence risk to include optimization of athletic performance and long-term athlete development. Interventions targeting movement quality, neuromuscular control, load tolerance, and recovery capacity may facilitate more robust adaptation to training demands and support safe and sustained sports participation during adolescence.

Several limitations should be acknowledged. This was a single-center retrospective study, and detailed information on sport type, competition level, training volume, and biological maturation could not be sufficiently analyzed. Because revisits were used as the outcome, mild cases that did not seek medical care may not have been captured. In addition, recurrence was determined based on visit history in medical records, and the exact timing of symptom resolution and changes in activity levels could not be directly assessed. Nevertheless, by focusing on clinically meaningful pain leading to medical revisits and by quantitatively and structurally visualizing recurrence patterns in adolescent athletes, this study provides important clinical insights into the dynamics of sports-related pain during growth.

Future studies should integrate sport-specific exposure, biological maturation indices, training load metrics, and biomechanical assessments to further elucidate mechanisms underlying inter-site recurrence dynamics. Prospective longitudinal designs may enable identification of predictive markers for vulnerability shifts within the apophyseal network and evaluation of targeted preventive interventions. Such efforts would facilitate development of personalized load-management strategies and contribute to a more comprehensive understanding of growth-related musculoskeletal adaptation in young athletes.

## Conclusion

This study is the first to objectively define revisit patterns for groin pain, anterior knee pain, and heel pain in adolescent athletes and to structurally visualize them as local and spatial recurrence. The results demonstrate that same-site recurrence is relatively uncommon, whereas pain frequently manifests as dynamic transitions across anatomically related sites. These findings provide a novel perspective that conceptualizes sports-related pain during growth not as isolated site-specific pathology but as a systems-level phenomenon based on an “apophyseal vulnerability network.” Clinically, the findings underscore the importance of moving beyond localized treatment toward comprehensive evaluation and proactive enhancement of global functional capacity to support recurrence prevention and sustained athletic performance.

## Data Availability

The datasets generated and/or analyzed during the current study are not publicly available due to institutional and ethical restrictions regarding patient confidentiality but are available from the corresponding author on reasonable request, subject to institutional approval.

## Acknowledgement

The authors thank the clinical and administrative staff of Ashiya Central Hospital for their support in data collection and clinical record management.

## References

1. Micheli LJ, Ireland ML. Prevention and management of calcaneal apophysitis in children. Clin Sports Med. 1987;6(2):337–346.

2. James AM, Williams CM, Haines TP. Effectiveness of treatment for calcaneal apophysitis: a systematic review. J Foot Ankle Res. 2013;6:16. doi:10.1186/1757-1146-6-16

3. Perhamre S, Janson S, Norlin R, Klässbo M. Sever’s injury: clinical diagnosis, treatment, and outcome. Scand J Med Sci Sports. 2011;21(6):e455–e460. doi:10.1111/j.1600-0838.2010.01201.x

4. Rathleff MS, Roos EM, Olesen JL, Rasmussen S. High prevalence and persistence of patellofemoral pain in adolescent females. Br J Sports Med. 2015;49(6):390–394. doi:10.1136/bjsports-2014-093684

5. Rathleff MS, Vicenzino B, van Middelkoop M, et al. Patellofemoral pain in adolescence and adulthood: same same, but different? Br J Sports Med. 2015;49(6):384–385. doi:10.1136/bjsports-2014-094245

6. van der Heijden RA, Lankhorst NE, van Linschoten R, Bierma-Zeinstra SMA, van Middelkoop M. Exercise therapy for adolescents and adults with patellofemoral pain syndrome. Br J Sports Med. 2015;49(14):927–934. doi:10.1136/bjsports-2014-093964

7. DiFiori JP, Benjamin HJ, Brenner JS, et al. Overuse injuries and burnout in youth sports: a position statement from the American Medical Society for Sports Medicine. Br J Sports Med. 2014;48(4):287–288. doi:10.1136/bjsports-2013-093299

8. Brenner JS. Council on Sports Medicine and Fitness. Sports specialization and intensive training in young athletes. Pediatrics. 2016;138(3):e20162148. doi:10.1542/peds.2016-2148

9. Jayanthi N, Pinkham C, Dugas L, Patrick B, LaBella C. Sports specialization in young athletes: evidence-based recommendations. Sports Health. 2013;5(3):251–257. doi:10.1177/1941738112464626

10. Baxter-Jones ADG, Eisenmann JC, Sherar LB. Controlling for maturation in pediatric exercise science. Pediatr Exerc Sci. 2005;17(1):18–30. doi:10.1123/pes.17.1.18

11. Benjamin M, McGonagle D. The anatomical basis for disease localization in entheses and related tissues. J Anat. 2001;199(Pt 5):503–526. doi:10.1046/j.1469-7580.2001.19950503.x

12. Hulme A, Finch CF, Nielsen RO. What is injury prevention? A systems-based approach to injury causation and prevention. Br J Sports Med. 2019;53(21):1384–1389. doi:10.1136/bjsports-2018-100059

